# Multimodal Prediction of Distressing and Persistent Psychotic-Like Experiences in Adolescence across Environment, Genetics, and Neuroimaging

**DOI:** 10.64898/2026.10.22.26365776

**Authors:** Abhay Lal, Nicole R. Karcher, Jonathan Ahern, Alison Rigby, Shefali Rai, Kristin Cadenhead, Janosch Linkersdörfer, Donald J. Hagler, Diliana Pecheva, Deanna M. Barch, Terry L. Jernigan, Pravesh Parekh, Anders M. Dale, Carolina Makowski

## Abstract

Adolescent psychotic-like experiences, particularly those that are distressing and persistent (dpPLEs), may elevate risk for a range of psychiatric disorders, yet the relative predictive contributions of brain structure, genetics, and environment to dpPLEs remain unknown. Using four timepoints from 7,399 children in the Adolescent Brain Cognitive Development (ABCD) Study, we developed stacked ensemble machine-learning models integrating cortical morphometry and whole-brain microstructural properties, polygenic risk scores, family psychiatric history, and childhood environmental factors to predict adolescent dpPLEs. Environmental predictors alone, driven by area deprivation index, neighborhood drug-related crime, and cumulative early adversity, were the strongest predictive domain (ROC-AUC = 0.687 ± 0.003), outperforming genetic (0.619 ± 0.020) and all imaging models (0.588-0.650). Adding genetic risk to environmental features produced the best-performing model overall (ROC-AUC = 0.697 ± 0.007), whereas adding neuroimaging yielded no further incremental gain (ROC-AUC=0.683-0.696). Beyond environmental exposures, family history of psychosis, cross-disorder polygenic risk, and both globally distributed and regionally specific imaging components contributed to prediction. Childhood environmental factors thus capture a core predictive signal preceding dpPLEs, while genetic and neuroimaging features added comparatively modest discriminative prediction value, but may offer complementary biological insights.

## INTRODUCTION

Psychotic-like experiences (PLEs), encompassing subclinical perceptual distortions, disorganized communication and unusual thought content, are reported in approximately 9% of U.S. adults, and 17-60% of youth experience some form of PLEs^1,2^. Although PLEs are relatively common in the general population, experiences that are both distressing and persistent across time may capture a subgroup of youth at higher risk for later mental health concerns^3,4^. This is supported by previous research identifying that youth with distressing persistent PLEs (dpPLEs) exhibit the most pronounced impairments across cognitive, neuroimaging, environmental, and functional domains compared to youth with either lower levels or non-distressing/transient PLEs^5^. DpPLEs also confer risk for outcomes in adulthood that span a transdiagnostic spectrum beyond psychotic disorders to encompass mood, anxiety, and behavioral conditions^4^, underscoring the broad clinical relevance of dpPLEs and the importance of studying premorbid factors early in development and before the emergence of dpPLEs.

Adolescent PLEs have been linked to genetic factors, with twin heritability estimates ranging from 15-59%^6^. Polygenic risk markers derived from large-scale genome-wide association studies (GWAS) for schizophrenia, bipolar disorder, and major depression have also been associated with PLEs in childhood^7^, adolescence^8^, and young adulthood^9^. This suggests that genetic vulnerability to adult-onset psychiatric disorders may already contribute to PLEs in youth^10,11^. Given the transdiagnostic predictors^12^ and outcomes^3,4,13,14^ associated with PLEs, recent GWAS highlighting shared genetic architecture across major psychiatric disorders^15–17^ may be particularly relevant to integrate as early genetic risk markers for dpPLEs.

Many of the genetic variants that shape mental health risk also shape brain structure^18,19^. Previous work has shown that brain structure is linked with polygenic risk for major psychiatric disorders and PLEs^7,20^. Beyond volume, differences between youth with dpPLEs and low levels of PLEs have been found in cortical surface area and thickness^21^, consistent with independent samples showing widespread structural abnormalities across frontal, temporal, and parietal regions in youth with psychosis-spectrum symptoms^22–24^. However, these analyses have relied predominantly on predefined regions of interest and traditional morphometric measures, potentially obscuring signals that traverse regional boundaries or reside in developmentally sensitive microstructural features^25^.

Neuroimaging-derived phenotypes that index microstructural tissue properties, particularly residing within deeper subcortical structures and peri-cortically, are dynamically changing during adolescence and may have developmental and theoretical relevance to dpPLE prediction^26–29^. They also may provide complementary, as well as distinct, neuroanatomical information relative to cortical morphometric measures such as cortical thickness, surface area, and sulcal depth. For instance, cortical gray-white contrast, measured from commonly acquired T1-weighted acquisitions and capturing patterns of intracortical myelin^26,30^, has been shown to capture case-control group differences in patients with psychotic disorders with greater sensitivity than other cortical metrics (e.g., cortical thickness)^26,31^, and tracks psychosis-related longitudinal outcomes^32^. Multi-shell diffusion models, such as restriction spectrum imaging^33^, also offer a valuable framework to measure developmentally sensitive changes in brain tissue microstructure, such as putative markers of cellular density, myelin, neuroinflammation, and glial morphology, which have been shown to change substantially across late childhood and early adolescence^27,28^. Measures of restricted or intracellular diffusion in particular can capture microstructural changes beyond commonly studied white matter tracts, including within subcortical structures and cortex, and may be meaningful predictors of early and dynamic brain structural changes in dpPLEs. These tissue microstructural properties have not yet been examined prior to emergence of adolescent dpPLEs.

Environmental context and adversity also substantially contribute to dpPLE emergence and play an important role in neurodevelopmental and mental health outcomes^14,34,35^. For example, urban residence is associated with a two- to three-fold increased risk of psychosis spectrum outcomes, likely driven by correlated exposures including deprivation, social fragmentation, and toxin exposure, and may also be relevant for dpPLEs^36,37^. Measures of urbanicity, area deprivation, poverty rate, and lead exposure risk have also been associated with PLEs in youth^38,39^. Cumulative stress, including discrimination and adverse childhood events, further compounds risk for PLE emergence^14^ which may impact brain circuitry important for emotional regulation and cognitive development^40^. Indeed, prior work has shown that brain volume mediates 11-25% of the association between environmental adversity and PLEs^38^, highlighting the importance of both neural and environmental pathways in shaping PLE risk. In the same vein, findings from a twin adolescent sample from the UK found that environmental and genetic factors interacted to further elevate risk for PLEs, emphasizing that these domains do not act in isolation^41^.

Despite evidence that brain structural patterns, genetic risk, and environmental factors are all associated with PLEs in adolescence, these three domains have not yet been examined together in a predictive framework. Large prospective studies, such as the Adolescent Brain Cognitive Development (ABCD) Study, offer a powerful lens to study such risk signatures prospectively across high-dimensional neuroimaging, genetic, and environmental factors. Here, we leverage baseline data from approximately 7000 participants in the ABCD Study (ages 9-10) to build an interpretable multimodal predictive framework for the emergence of later dpPLEs in adolescence through three aims: (1) to test whether later dpPLE risk is preceded by a detectable baseline vulnerability signature across multimodal measures; (2) to compare the predictive signal contributed by whole-brain structural MRI, diffusion MRI, genetic risk, and environmental exposures; (3) to identify imaging and non-imaging features that consistently drive model predictions. We hypothesized that complementary, multimodal integration would improve prediction beyond any single domain, and that the novel integration of high-dimensional cortical anatomy and whole-brain tissue microstructure may elucidate neurobiological patterns that may be most relevant for prediction of dpPLEs and other mental health concerns.

## METHODS

### Participants

Data were drawn from the ABCD Study^42^, a prospective, multisite, longitudinal investigation of child brain development and health that enrolled 11,878 children aged 9-10 years across 21 U.S. sites. The ABCD Study includes data collection spanning neuroimaging, genetics, environmental and geocoded exposures, physical and mental health, cognition, and family history, with participants assessed at baseline and at regular follow-up visits. The design and recruitment details of the ABCD Study have been described previously^43,44^.

### Identifying youth with dpPLEs

Psychotic-like experiences were assessed annually from baseline through 6-year follow-ups using the Prodromal Questionnaire-Brief Child Version (PQ-BC), a 21-item self-report measure validated for use in this age range^2^. For each endorsed item, children reported associated distress, yielding both a total symptom score and a distress-weighted score at each visit. Consistent with prior ABCD work^5^, participants were classified into the dpPLE group (n=322) if their PQ-BC distress score was ≥ 1.96 standard deviations (SDs) above the mean for at least two of their last three available waves of data. Individuals with low PQ-BC severity scores ≤ 0.50 SDs below the mean across the three last available waves were classified into the low PLE group (n=7,077). Given that we were interested in evaluating features that were most predictive of dpPLEs compared to having a low level of PLEs, participants with transient or non-distressing PLEs (n=1731) were excluded from the binary outcome (see Supplementary Materials for details on participant inclusion/exclusion).

### Predictor domains

Baseline predictors across neuroimaging, genetic, and environmental domains are described in Supplementary Table 1, with a schematic of all imaging features provided in Supplementary Figure 1.

#### Neuroimaging features

Imaging predictors comprised vertex-wise structural MRI (sMRI) measures and voxel-wise diffusion MRI (dMRI) measures, processed through the standardized ABCD imaging pipeline^45^. Vertex-wise structural features included cortical surface area, sulcal depth, gray-white contrast (GWC), and cortical thickness. Voxel-wise diffusion features were derived from restriction spectrum imaging (RSI) model fit to diffusion weighted imaging and consisted of restricted normalized directional (RND) and restricted normalized isotropic (RNI) signal fractions^46^. Modality-specific quality control inclusion flags were applied before analysis as recommended by the ABCD Study Consortium^26^.

#### Environmental features

Environmental predictors captured household economic hardship, neighborhood and residential context, and exposure to adverse life events. Financial adversity was operationalized as a summed score across seven caregiver-reported indicators of material and economic hardship. Neighborhood safety was indexed using three caregiver-reported items. Geocoded contextual measures were derived from external data linked to participants’ residential locations, as described in previous work^47,48^. These neighborhood-based factors included years at current residence, drug-related neighborhood crime, national percentile on the Area Deprivation Index (ADI), estimated lead-exposure risk, and neighborhood poverty rate. Adverse life events were summarized as a cumulative count across 33 candidate events, counting only events endorsed as experienced, appraised as negative, and reported to have affected the child at least somewhat. In total, eight environmental predictors were derived from 148 unique variables (Supplementary Table 1).

#### Genetic and family history features

The genetic domain comprised family history variables and polygenic scores (PGS). Family history of psychopathology was derived from caregiver reports of relevant behaviors in any blood relative. Included features were indicators of any psychiatric history regardless of degree of relatedness, and of having a first-degree relative that had experienced depression, mania, hallucinations, and/or delusions. GWAS summary statistics for schizophrenia^49^, major depression^50^, and bipolar disorder^51^, and a cross-disorder psychopathology score (P-factor)^15^ aggregating shared liability across 14 psychiatric disorders were leveraged to calculate PGS using PGScs^52,53^. GWAS sample details are included in Supplementary Table 2. PGS derived from predominantly European-ancestry GWAS show reduced portability to multi-ancestry samples; thus, we additionally compared prediction of dpPLEs from PGS derived in European and non-European subsamples separately. See Supplementary Materials for more details of PGS calculation and genomic ancestry estimation.

### Overview of predictive modeling

We trained interpretable machine learning models to predict dpPLE or lowPLE outcomes from each predictor domain individually and from multimodal combinations, following recommended practices in machine learning models in psychiatry^54^. We used Shapley Additive exPlanations (SHAP)^55^ to quantify the contribution of individual features and modalities to model predictions. All analyses were implemented in Python 3 using scikit-learn^56^; base and meta-level classifiers were regularized logistic regression models. The full analytic workflow is shown in **Figure 1** and full implementation details provided in Supplementary Materials.

**Figure 1.**
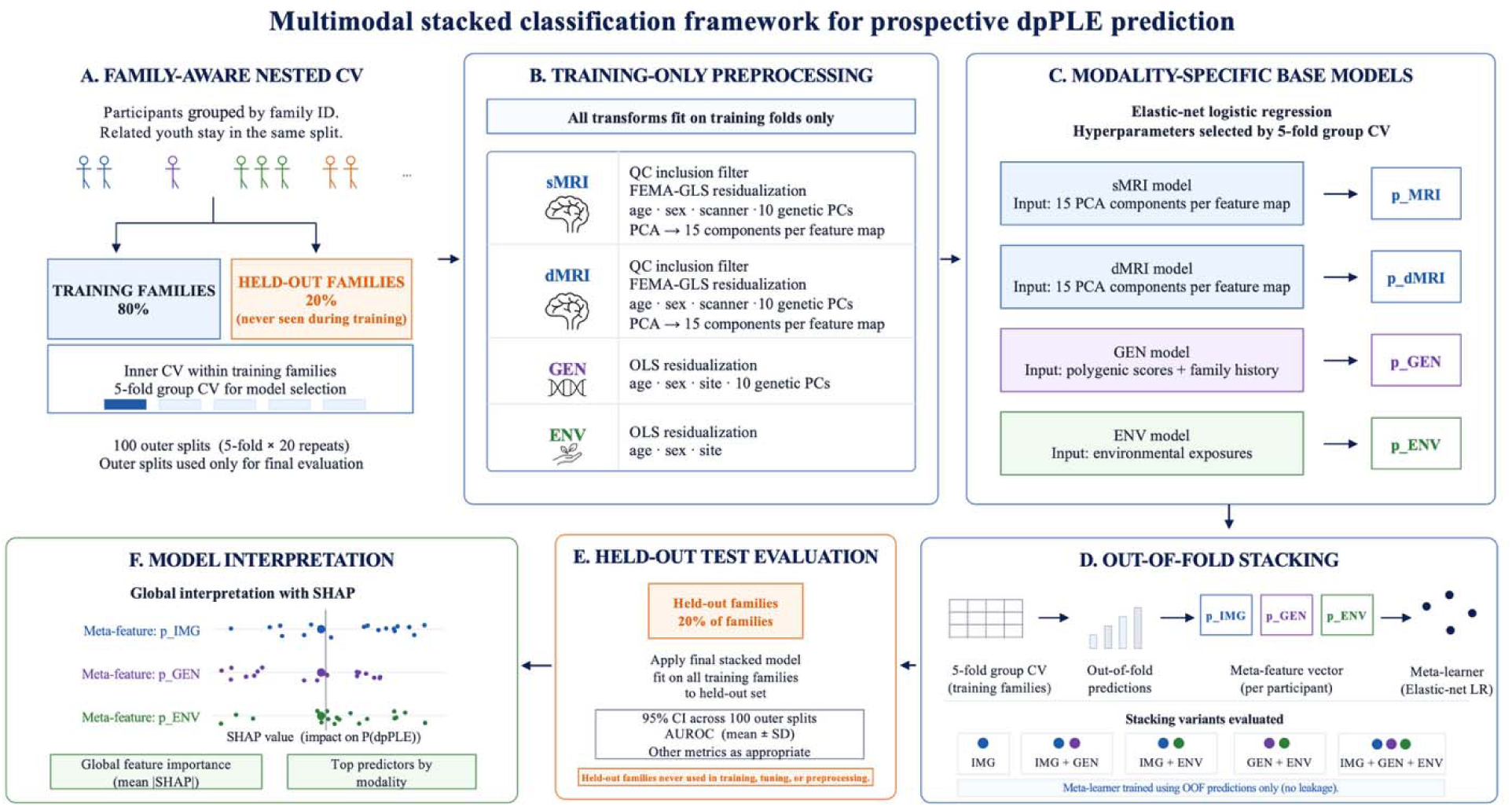
Multimodal stacked classification framework for predicting dpPLEs. A. Family-aware nested cross-validation. Participants were grouped by family so that siblings were never split across training and test sets. Performance was estimated across 100 outer splits (20 repeats of 5-fold stratified cross-validation), with hyperparameters selected in a 5-fold inner loop within each outer training fold. B. Training-only preprocessing. All transformations were fit on the training fold and applied unchanged to the test fold. Vertex-wise sMRI and voxel-wise dMRI maps were residualized for age, sex, scanner, and the first ten genetic ancestry PCs using FEMA generalized least squares with family as a random effect, then standardized and reduced to 15 PCs per feature map (for example, cortical thickness, surface area for sMRI; RNI, RND for dMRI). GEN and ENV predictors were residualized by ordinary least squares. C. Modality-specific base models. Separate regularized logistic regression models were trained for each imaging feature map (15 PCs each) and for GEN and ENV predictors, each returning a predicted probability of dpPLE membership. D. Out-of-fold stacking. Base-model predictions for training participants were generated out-of-fold through a further level of family-grouped 5-fold cross-validation and used as meta-features for a regularized logistic regression meta-learner. Five configurations of imaging/genetic/environmental predictive domains were evaluated. E. Held-out evaluation. ROC-AUC and PR-AUC were computed in each held-out test fold, averaged across the five folds of each repeat, and summarized as mean ± SD across the 20 repeats. F. Model interpretation. SHAP values quantified the contribution of each meta-feature to prediction of dpPLEs and of each PC to imaging base-model predictions, with importance summarized as mean |SHAP|. Abbreviations: ENV, environmental; GEN, genetic and family history; IMG, imaging (a single sMRI or dMRI feature map); PC, principal component; PR-AUC, area under the precision-recall curve; ROC-AUC, area under the receiver operating characteristic curve; SHAP, SHapley Additive exPlanations.

### Cross-validation framework

Predictive performance was estimated using repeated, family-based nested cross-validation (**Figure 1A**). We performed 20 repeats of 5-fold stratified cross-validation, yielding 100 outer splits, with hyperparameters selected in a 5-fold inner loop nested within each outer training fold, so that performance was always estimated on data unseen during both model fitting and model selection. Stratification preserved the proportion of dpPLE to lowPLE cases within folds. Family identifiers were used as grouping variables so that siblings were never split across training and test sets, preventing leakage from shared genetic and common environmental factors within a family^57,58^.

### Preprocessing of input features

All predictors were preprocessed within the cross-validation framework, with every parameter estimated on the training fold only and applied unchanged to the held-out test fold (**Figure 1B**). Vertex-wise sMRI and voxel-wise dMRI maps were residualized for baseline age, sex, study site, scanner, and the first ten genetic ancestry principal components using a generalized least-squares estimator implemented in the Fast and Efficient Mixed-effects Algorithm (FEMA)^55^, with family as a random effect, then standardized and reduced by principal component analysis (PCA) to retain 15 components per feature map. Non-imaging predictors were residualized by ordinary least squares regression: genetic and family history features accounted for baseline age, sex, study site, and the first ten genetic principal components, and environmental features accounted for baseline age, sex, and study site. Full preprocessing parameters are provided in Supplementary Materials.

### Base-model development

Separate base models were trained for each predictor domain, including one model per imaging feature map (e.g., surface area, sulcal depth, GWC, cortical thickness for sMRI; RNI and RND for dMRI), each taking 15 principal components as input (**Figure 1C**). All base models were regularized logistic regression classifiers. The penalty type (L1, L2, or elastic net), regularization strength, L1 ratio (for elastic net) and class weighting were selected by grid search in the inner 5-fold cross-validation loop, and the best configuration was refit on the full training fold. Solver settings, the hyperparameter grid, and model-selection metrics are provided in Supplementary Materials.

### Stacked multimodal models

To integrate information across domains, we used stacked generalization ^59,60^ (**Figure 1D**). Stacked generalization is a powerful method that uses a meta-learner to combine the predictions of multiple base models, and has shown success as a classification tool in various biomedical signal processing contexts ^61^. Each base model returned a single predicted probability of dpPLE group membership per participant, and these predicted probabilities (hereafter meta-features) were treated as predictors in a further regularized logistic regression (the meta-learner) that learned how much weight each domain should carry. Stacking was preferred over fitting a single logistic regression to the pooled feature set because the domains differed substantially in dimensionality and signal density (15 imaging principal components against 8 genetic and 8 environmental predictors) and stacking allowed each domain to be tuned to its own optimal penalty type and strength. To prevent leakage, meta-features for training participants were generated out-of-fold using a further level of 5-fold family-grouped stratified cross-validation within each outer training fold. Meta-features for the outer test fold were generated from base models refitted on the full outer training fold. The meta-learner was tuned over the same grid as the base models.

Five configurations were evaluated: (i) imaging only; (ii) imaging plus genetic; (iii) imaging plus environmental; (iv) non-imaging only (genetic plus environmental); and (v) a full multimodal model combining all three domains. All configurations were fit on identical participants and evaluated on identical outer test folds, so differences in performance are attributable only to which domains were available. Configuration (i) supplied a single meta-feature and therefore was equivalent to a recalibration of the imaging base model.

### Prediction performance metrics

Performance was evaluated on the held-out outer test folds (**Figure 1E**). The receiver operating characteristic - area under the curve (ROC-AUC) and precision recall AUC (PR-AUC) were computed within each held-out test fold, averaged across the five folds of each repeat, and reported as the mean ± SD of the resulting 20 repeat-level estimates. Because the five test folds within a repeat are mutually exclusive and exhaustive, every participant contributed exactly once to each repeat-level estimate. The reported SD should therefore be interpreted as the stability of the performance estimate under repeated random partitioning of this sample, rather than as a confidence interval for generalization to a new sample.

### Model interpretability (SHAP)

Feature contributions were quantified with SHAP^55^, computed within each outer split for held-out participants only (**Figure 1F**). For imaging-only models, fitting occurred in PCA space, so SHAP values indexed the contribution of the 15 principal components retained within each training fold rather than individual vertices or voxels. For stacked models, SHAP values quantified the contribution of each base-model prediction to the meta-model output, placing imaging, genetic, and environmental information on a common scale. Feature contributions are reported as the mean absolute SHAP value per feature, computed within each held-out test fold, averaged across the five folds of each repeat, and summarized as the mean ± SD of the resulting 20 repeat-level estimates, matching the aggregation used for performance metrics. For each imaging feature map, the loadings of the component with the highest mean absolute SHAP value were displayed on the brain alongside its component-level SHAP values, reflecting descriptive maps of what the leading component encodes spatially rather than voxelwise importance maps.

## RESULTS

### Participants

A total of 322 participants meeting criteria for dpPLEs and 7,077 individuals with a low level of PLEs were included in the analysis. See **Table 1** for demographics and descriptive statistics for severity scores drawn from the Prodromal Questionnaire-Brief Child Version (PQ-BC) and non-imaging features input into the prediction model.

**Table 1.** Participants and descriptive statistics. Statistical tests compare the dpPLE and low PLE groups. Continuous variables are summarized as mean ± SD and compared with a two-tailed Welch t-test. Categorical variables are compared with an omnibus chi-square test on the 2 × k table, reported once per variable on the section row. Skewed count variables are summarized as median [IQR] and compared with a two-tailed Mann–Whitney U test. Effect sizes: d = Cohen’s d; V = Cramer’s V; r = rank-biserial correlation; OR = odds ratio with 95% CI for the second listed category (2 × 2 tables only). Positive d and r indicate higher values in dpPLE. P values are exact and unadjusted for multiple comparisons. Abbreviations. PLE, psychotic-like experiences; dpPLE, distressing and persistent psychotic-like experiences; sMRI, structural magnetic resonance imaging; dMRI, diffusion magnetic resonance imaging; SD, standard deviation; HS, high school; GED, general educational development; PGS, polygenic scores; IQR, interquartile range; ADI, area deprivation index.

| Characteristic | dpPLE group<br>(n = 322) | Low PLE group<br>(n = 7,077) | dpPLE vs Low PLE | Effect size |
| --- | --- | --- | --- | --- |
| **N, Age, Sex** |  |  |  |  |
| N (sMRI / dMRI) | 308 / 279 | 6,781 / 6,157 |  |  |
| Baseline age (mean ± SD) | 9.86 ± 0.62 | 9.97 ± 0.63 | $t(351) = -3.17, P = 0.002$ | $d = -0.18$ |
| Sex Male / Female | 91 / 231 | 4,038 / 3,039 | $\chi^2(1) = 102.39, P = 4.6 \times 10^{-24}$ | OR = 3.37<br>[2.63, 4.32] |
| **Race / Ethnicity** | | | $\chi^2(4) = 56.93, P = 1.3 \times 10^{-11}$ | **V = 0.088** |
| White non-Hispanic | 132 (41.0%) | 4,162 (58.8%) |  |  |
| Black / African American | 71 (22.0%) | 813 (11.5%) |  |  |
| Hispanic | 81 (25.2%) | 1,262 (17.8%) |  |  |
| Asian | 3 (0.9%) | 144 (2.0%) |  |  |
| Other / Multiracial | 35 (10.9%) | 693 (9.8%) |  |  |
| **Household Income** | | | $\chi^2(2) = 103.77, P = 2.9 \times 10^{-23}$ | **V = 0.123** |
| < \$50k | 139 (48.9%) | 1,560 (23.7%) | | |
| \$50k – \$100k | 78 (27.5%) | 1,901 (28.9%) | | |
| > \$100k | 67 (23.6%) | 3,125 (47.4%) | | |
| **Highest Parental Education** | | | $\chi^2(4) = 88.57, P = 2.6 \times 10^{-18}$ | **V = 0.110** |
| < HS diploma | 25 (7.8%) | 235 (3.3%) |  |  |
| HS diploma / GED | 37 (11.6%) | 518 (7.4%) |  |  |
| Some college | 125 (39.1%) | 1,610 (22.9%) |  |  |
| Bachelor's degree | 63 (19.7%) | 1,908 (27.1%) |  |  |
| Post-graduate degree | 70 (21.9%) | 2,758 (39.2%) |  |  |
| **PQ-BC Severity Score (mean ± SD)** |  |  |  |  |
| Baseline | 16.13 ± 17.47 | 4.46 ± 8.46 | $t(327) = 11.90, P = 2.3 \times 10^{-27}$ | $d = +1.29$ |
| Year 1 | 16.80 ± 18.86 | 2.66 ± 6.54 | $t(307) = 13.07, P = 2.5 \times 10^{-31}$ | $d = +1.89$ |
| Year 2 | 17.96 ± 16.57 | 1.68 ± 4.34 | $t(309) = 17.22, P = 4.7 \times 10^{-47}$ | $d = +2.99$ |
| Year 3 | 18.24 ± 14.54 | 0.94 ± 2.57 | $t(287) = 20.15, P = 7.5 \times 10^{-57}$ | $d = +4.47$ |
| Year 4 | 24.75 ± 16.50 | 0.60 ± 1.30 | $t(266) = 23.91, P = 3.2 \times 10^{-68}$ | $d = +6.74$ |
| Year 5 | 23.64 ± 14.80 | 0.47 ± 1.10 | $t(249) = 24.75, P = 4.3 \times 10^{-69}$ | $d = +7.24$ |
| Year 6 | 16.41 ± 15.55 | 0.54 ± 2.01 | t(257) = 16.39, P = 7.8 × 10<sup>-42</sup> | d = +4.27 |
| **Polygenic Scores (mean ± SD)** |  |  |  |  |
| SCZ PGS | 2.90e-07 ± 1.61e-07 | 2.31e-07 ± 1.62e-07 | t(351) = 6.48, P = 3.0 × 10<sup>-10</sup> | d = +0.37 |
| DEP PGS | 2.86e-08 ± 3.18e-08 | 1.25e-08 ± 3.43e-08 | t(356) = 8.89, P = 3.2 × 10<sup>-17</sup> | d = +0.47 |
| BIP PGS | 9.71e-08 ± 3.60e-08 | 8.49e-08 ± 3.74e-08 | t(353) = 5.92, P = 7.7 × 10<sup>-9</sup> | d = +0.33 |
| Cross-disorder PGS | -4.82e-08 ± 9.39e-08 | -9.21e-08 ± 9.78e-08 | t(352) = 8.17, P = 5.5 × 10<sup>-15</sup> | d = +0.45 |
| **Family History (Yes / No, % positive)** |  |  |  |  |
| Depression (1st degree) | 184 / 138 (57.1%) | 3,125 / 3,952 (44.2%) | χ<sup>2</sup>(1) = 20.49, P = 6.0 × 10<sup>-6</sup> | OR = 1.69<br>[1.35, 2.11] |
| Mania (1st degree) | 49 / 273 (15.2%) | 592 / 6,485 (8.4%) | χ<sup>2</sup>(1) = 17.42, P = 3.0 × 10<sup>-5</sup> | OR = 1.97<br>[1.43, 2.70] |
| Hallucinations/Delusions (1st degree) § | 72 / 250 (22.4%) | 818 / 6,259 (11.6%) | χ<sup>2</sup>(1) = 32.95, P = 9.5 × 10<sup>-9</sup> | OR = 2.20<br>[1.68, 2.89] |
| Hallucinations/Delusions (any family history) § | 33 / 289 (10.2%) | 386 / 6,691 (5.5%) | χ<sup>2</sup>(1) = 12.37, P = 4.4 × 10<sup>-4</sup> | OR = 1.98<br>[1.36, 2.88] |
| **Environmental Predictors** |  |  |  |  |
| Adverse life events count (median [IQR]) ‡ | 0 [0–2]; 47.5% ≥ 1 | 0 [0–1]; 32.8% ≥ 1 | U = 1,332,475, P = 7.2 × 10<sup>-10</sup> | r = +0.17 |
| Financial adversity count (median [IQR]) | 0 [0–1]; 38.2% ≥ 1 | 0 [0–0]; 17.3% ≥ 1 | U = 1,377,946, P = 3.0 × 10<sup>-21</sup> | r = +0.21 |
| Neighborhood safety score (mean ± SD) | 2.40 ± 1.03 | 2.03 ± 0.93 | t(345) = 6.30, P = 9.1 × 10<sup>-10</sup> | d = +0.39 |
| Years at current residence (mean ± SD) | 4.84 ± 3.66 | 5.70 ± 3.76 | t(352) = -4.09, P = 5.3 × 10<sup>-5</sup> | d = -0.23 |
| Drug-related crime count (median [IQR]) | 3,958 [1,215–8,005] | 2,694 [841–6,574] | U = 1,341,708, P = 6.6 × 10<sup>-8</sup> | r = +0.18 |
| ADI national percentile (mean ± SD) | 48.20 ± 28.43 | 37.47 ± 25.68 | t(345) = 6.65, P = 1.2 × 10<sup>-10</sup> | d = +0.42 |
| Lead exposure risk index (mean ± SD) | 5.86 ± 3.17 | 4.88 ± 3.04 | t(349) = 5.45, P = 9.5 × 10<sup>-8</sup> | d = +0.32 |
| Neighborhood poverty rate (median [IQR]) | 19.4 [10.8–33.6] | 13.0 [7.3–23.3] | U = 1,426,563, P = 1.9 × 10<sup>-14</sup> | r = +0.25 |

### Single-domain prediction performance

The environmental model alone achieved the highest prediction performance, with an ROC-AUC of 0.687 ± 0.003 (PR-AUC = 0.091 ± 0.001) (**Figure 2A**; Supplementary Table 3). This exceeded every individual imaging modality by an AUC of 0.037-0.099, and exceeded the genetic model (ROC-AUC = 0.619 ± 0.020; PR-AUC = 0.080 ± 0.006) by 0.068. Notably, the genetic domain, comprising polygenic scores and family psychiatric history, performed comparably to the best-performing vertex-wise cortical measure (both ROC-AUC = 0.619). Note that performance of non-imaging domains was driven largely by participants of predominantly European ancestry (environment: ROC-AUC= 0.712 ± 0.044; genetics: 0.679 ± 0.054) compared to participants of non-European-like ancestry (environment: 0.593 ± 0.046; genetics: 0.573 ± 0.050) (Supplementary Table 4).

**Figure 2.**
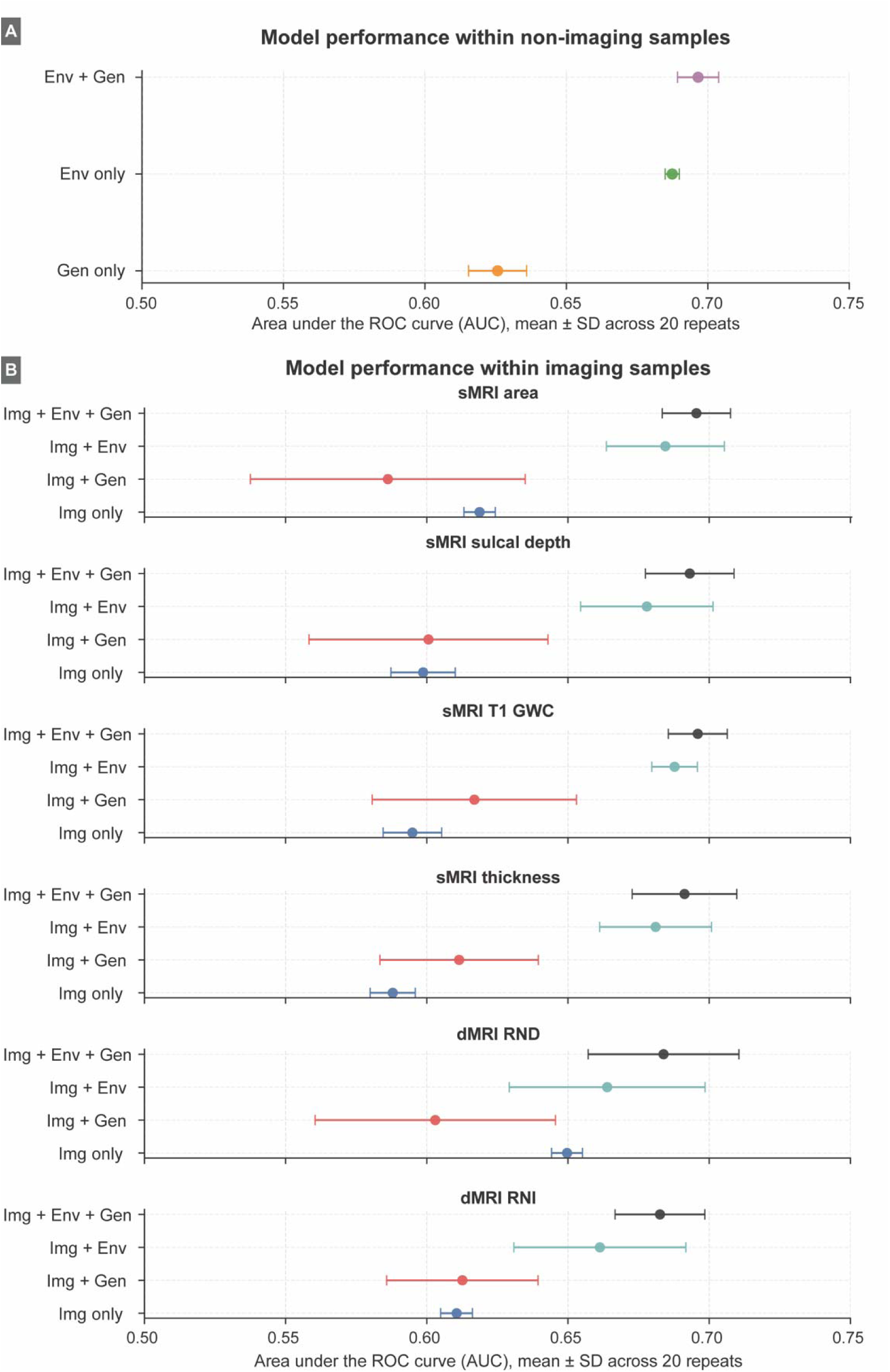
Prediction performance results. (A) ROC-AUC for each modality-specific imaging model (sMRI cortical area, sulcal depth, GWC, cortical thickness; dMRI RND, RNI) across four feature-set configurations (imaging only; imaging + environmental; imaging + genetic; imaging + environmental + genetic). Error bars represent ±1 SD across 20 repeats (100 total cross-validation splits: 5 folds × 20 repeats). (B) ROC-AUC for the non-imaging domain models (environmental only, genetic only, and environmental + genetic), plotted in the same format. Abbreviations. Env, environmental; Gen, genetic and family history; Img, imaging. sMRI, structural magnetic resonance imaging; GWC, gray-white contrast; dMRI, diffusion magnetic resonance imaging; RND, restricted normalized directional diffusion; RNI, restricted normalized isotropic diffusion; SD, standard deviation.

Across the six imaging measures (e.g. four cortical sMRI measures and two dMRI measures), the prediction of adolescent dpPLEs was modest (ROC-AUC range 0.588-0.650; PR-AUC 0.065-0.087; **Figure 2B**; Supplementary Table 5). Voxel-wise microstructural measures outperformed vertex-wise cortical measures: restricted normalized directional diffusion (RND) was the single best-performing imaging modality (ROC-AUC = 0.650 ± 0.005; PR-AUC = 0.087 ± 0.004), followed by restricted normalized isotropic (RNI; ROC-AUC = 0.611 ± 0.006; PR-AUC = 0.082 ± 0.003). Among the vertex-wise cortical measures, surface area performed best (ROC-AUC = 0.619 ± 0.006), followed by sulcal depth (0.599 ± 0.011), T1-weighted gray-white matter contrast (0.595 ± 0.010) and cortical thickness (0.588 ± 0.008). Repeat-level variability was low for all imaging models (SD ≤ 0.011), indicating stable but limited discrimination. Absolute PR-AUC values remained low across all models, consistent with the low base rate of dpPLEs in the cohort.

### Stacked multimodal prediction performance

We next tested which combination of domains improved prediction through stacked ensembles. Combining the two non-imaging domains yielded the highest performance observed in any model configuration (Env + Gen: ROC-AUC = 0.697 ± 0.007; PR-AUC = 0.100 ± 0.006), a gain of 0.010 AUC and 0.009 PR-AUC over the environmental model alone (a ∼10% relative improvement in PR-AUC) (**Figure 2**; Supplementary Table 6). Full multimodal models (Img + Env + Gen) achieved ROC-AUCs of 0.683-0.696 and PR-AUCs of 0.096-0.101. The best-discriminating configurations incorporated T1-weighted GWC (ROC-AUC = 0.696 ± 0.010) and cortical surface area (0.695 ± 0.012), while the highest PR-AUC was obtained with RNI (0.101 ± 0.007) (**Figure 2**; Supplementary Table 6).

No full multimodal configuration exceeded the non-imaging Env + Gen model (ROC-AUC = 0.697 ± 0.007; Supplementary Table 6), with differences ranging from -0.001 to -0.014 AUC. The same pattern held for two-domain stacks: Img + Env models (0.661-0.688) did not improve on the environmental model alone (0.687), and Img + Gen models (0.586-0.617) did not improve on the genetic model alone (0.619). Within every imaging modality, performance ordered consistently as Img + Env + Gen > Img + Env > Img + Gen, mirroring the standalone ordering of the non-imaging domains and indicating that ensemble performance was driven predominantly by the environmental domain, with a smaller genetic contribution and negligible incremental contribution from imaging.

Stacked models incorporating imaging also showed substantially greater variability across repeats (SD = 0.008-0.049) than non-imaging models (SD = 0.003-0.020), with the largest variability in Img + Gen configurations (e.g. sMRI area, SD = 0.049). This inflation was consistent with the smaller imaging QC subsample available for stacking and suggests that apparent differences between imaging-based stacks should be interpreted cautiously.

### Feature importance: SHAP analysis

To characterize which predictors drove model output, we computed mean absolute SHAP values in log-odds units for each model. These quantify the magnitude of a feature’s contribution to the predicted log-odds.

#### Environmental features

In the environmental model (**Figure 3**; Supplementary Table 7), the strongest contributors were indices of neighborhood disadvantage and early adversity: area deprivation index at the primary residential address (national percentile; 0.193 ± 0.013), neighborhood drug-related crime count (0.161 ± 0.013), and cumulative count of adverse life events (0.144 ± 0.005). Attributions were tightly distributed across repeats (SD ≤ 0.013), indicating a stable and consistently ranked set of environmental predictors. The two leading features index the socio-spatial context of the child’s residence rather than individual-level exposure, suggesting that neighborhood-level deprivation and crime may carry predictive information beyond directly experienced adversity.

**Figure 3.**
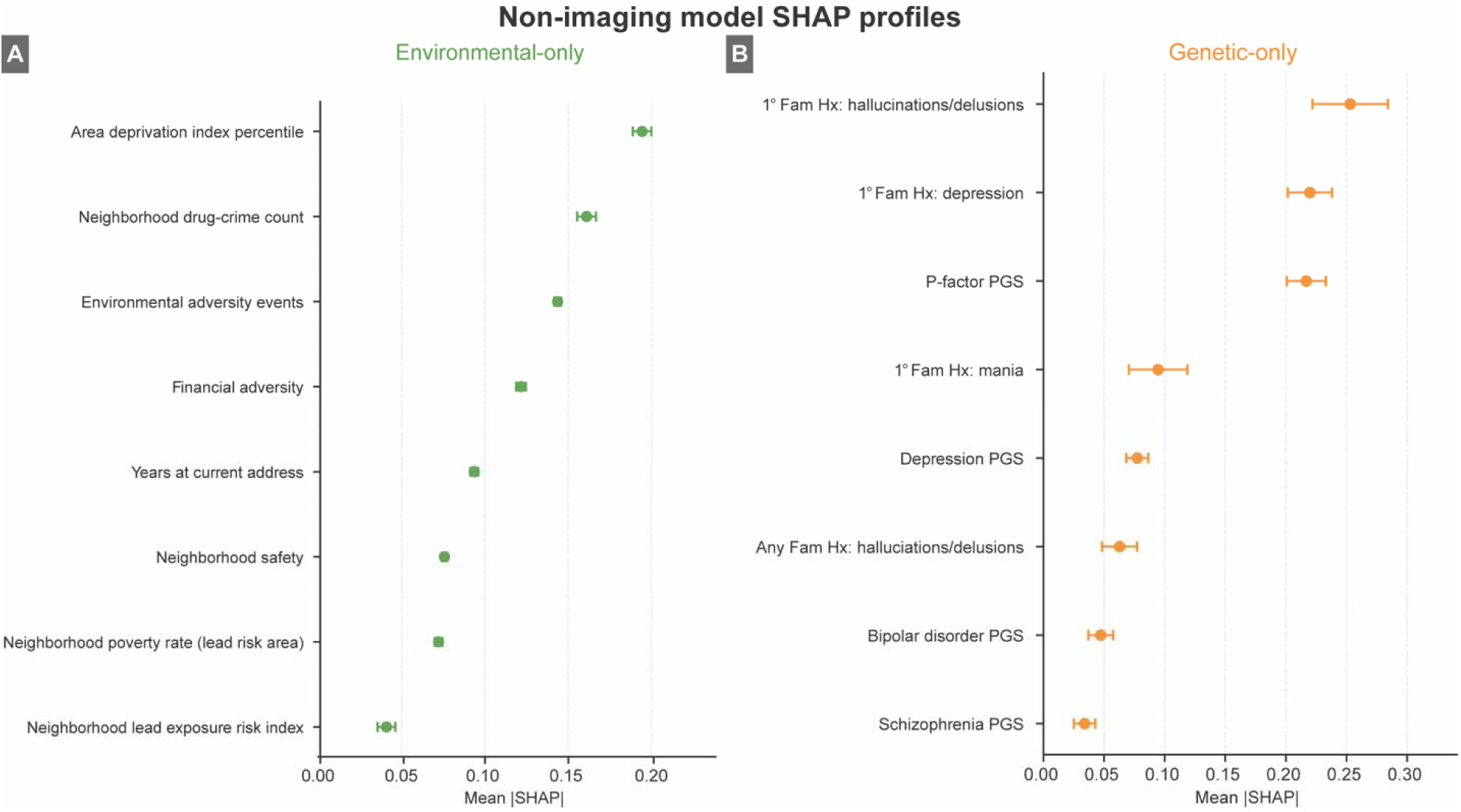
SHAP feature importance for non-imaging predictor domains. (A) Mean |SHAP| (± SD across 20 repeats) for the top-ranked environmental predictors, plotted as point estimates with horizontal error bars and ordered by mean |SHAP|. (B) Mean |SHAP| (± SD across 20 repeats) for the top-ranked genetic and family-history predictors, plotted in the same format. Abbreviations: ADI, Area Deprivation Index; Fam Hx, family history; PGS, polygenic score; SHAP, Shapley Additive exPlanations.

#### Genetic and family history features

In the genetic model (**Figure 3**; Supplementary Table 7), familial psychiatric history outweighed PGS predictors. First-degree family history of psychosis was the leading contributor (0.253 ± 0.071), followed by first-degree family history of depression (0.220 ± 0.042) and the p-factor PGS (0.217 ± 0.037). Attribution magnitudes for the top three features were closely spaced and their SDs comparatively wide, indicating that no single genetic-domain predictor dominated the predictive signal and that their relative ranking varied across repeats.

#### Imaging features

Within imaging-only models (Supplementary Table 8), attributions differed markedly in magnitude between voxel-wise diffusion (**Figure 4**) and vertex-wise cortical features (**Figure 5**). Models built on PCs of the voxel-wise dMRI measures yielded attributions an order of magnitude larger than those built on components of the vertex-wise cortical measures. For RND, the leading components were PC3 (SHAP values 3.858 ± 0.434), PC11 (2.116 ± 0.222) and PC9 (2.054 ± 0.127); for RNI, PC9 (2.110 ± 0.215), PC4 (1.667 ± 0.167) and PC7 (1.029 ± 0.114) (**Figure 4**). These findings support global microstructural properties as the most informative imaging signal for dpPLEs, particularly for directed diffusion within white matter tracts and isotropic diffusion within subcortical structures.

**Figure 4.**
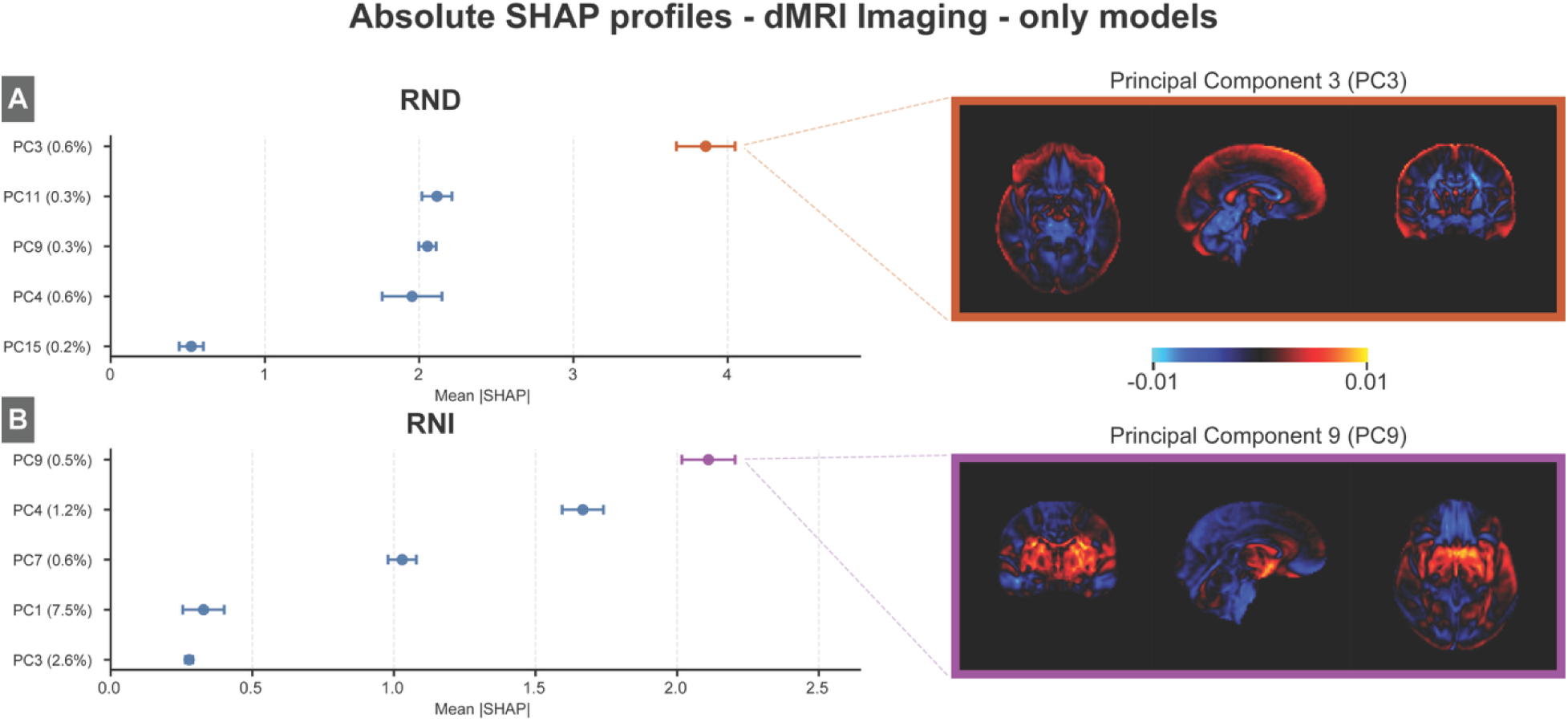
SHAP feature importance and back-projected principal-component (PC) brain maps for the voxel-wise diffusion imaging-only models. Absolute SHAP profiles (mean |SHAP| ± SD across 20 repeats) are shown for the top-ranked PCs of each dMRI metric (RND, RNI) plotted as point estimates with horizontal error bars and ordered by mean |SHAP|. For each metric, the loadings of the highest-ranked PC are back-projected to voxel space, where the color scale indicates magnitude of loadings. Abbreviations. dMRI, diffusion magnetic resonance imaging; PC, principal component; RND, restricted normalized directional diffusion; RNI, restricted normalized isotropic diffusion; SHAP, Shapley Additive exPlanations.

**Figure 5.**
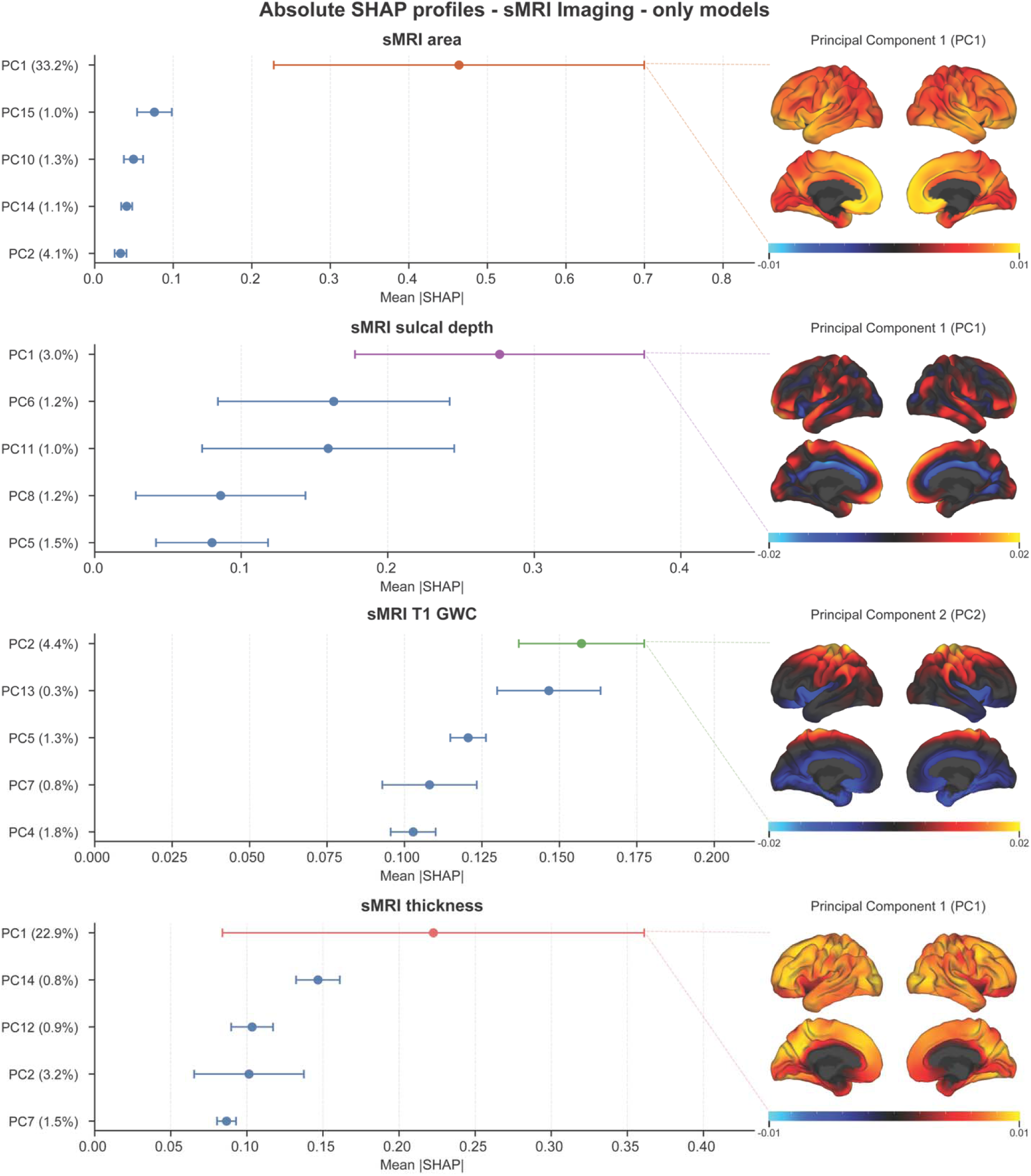
SHAP feature importance and back-projected principal-component (PC) brain maps for the vertex-wise cortical imaging-only models. Absolute SHAP profiles (mean |SHAP| ± SD across 20 repeats) are shown for the top-ranked PCs of each sMRI metric (cortical area, sulcal depth, T1 gray-white contrast, cortical thickness), plotted as point estimates with horizontal error bars and ordered by mean |SHAP|. For each metric, the loadings of the highest-ranked PC are back-projected to the inflated cortical surface (lateral and medial views, left and right hemispheres), where color scale indicates magnitude of loadings. Large SDs relative to means for the leading cortical components (e.g., for PC1 of cortical surface area) indicated that these attributions were unstable across repeats and thus should be interpreted with caution. Abbreviations. GWC, gray-white contrast; PC, principal component; SHAP, Shapley Additive exPlanations; sMRI, structural magnetic resonance imaging.

Driving features of vertex-wise cortical maps encompassed both global and regional brain patterns (**Figure 5**, Supplementary Table 8). Cortical surface area (PC1 = 0.464 ± 0.538), sulcal depth (PC1 = 0.276 ± 0.225) and cortical thickness (PC1 = 0.223 ± 0.316) were each led by the first principal component, which captures globally distributed variance, with substantially smaller contributions from higher-order, regionally specific components (e.g. area PC15 = 0.076 ± 0.050; thickness PC14 = 0.147 ± 0.033). T1-weighted gray-white matter contrast was the exception, distributing importance more evenly across mid-order components (PC2 = 0.157 ± 0.046; PC13 = 0.147 ± 0.038; PC5 = 0.121 ± 0.013), with the highest contribution from PC2 capturing positive loadings within sensori-motor cortex. Large SDs relative to means for the leading cortical components (e.g., for PC1 of cortical surface area) indicated that these attributions were unstable across repeats and thus should be interpreted with caution.

### Stacked model contributions

At the ensemble level, SHAP values were computed on the out-of-fold domain scores entering each stacked model, quantifying each domain’s contribution to the ensemble’s predicted log-odds (**Figure 6**). In the non-imaging stacked model, the environmental score contributed more than twice as much as the genetic score (ENV LR = 0.427 ± 0.124 vs GEN LR = 0.200 ± 0.107; Supplementary Table 9). This dominance persisted in stacks built on the vertex-wise cortical measures (Supplementary Table 9): in all four full multimodal sMRI configurations, the environmental score was the leading contributor (0.413-0.440), followed by the genetic score (0.209-0.249), with the imaging score contributing least (0.123-0.202). The single exception among these was the Img + Gen stack for cortical thickness, in which the imaging score marginally exceeded the genetic score (0.267 ± 0.151 vs 0.237 ± 0.075).

**Figure 6.**
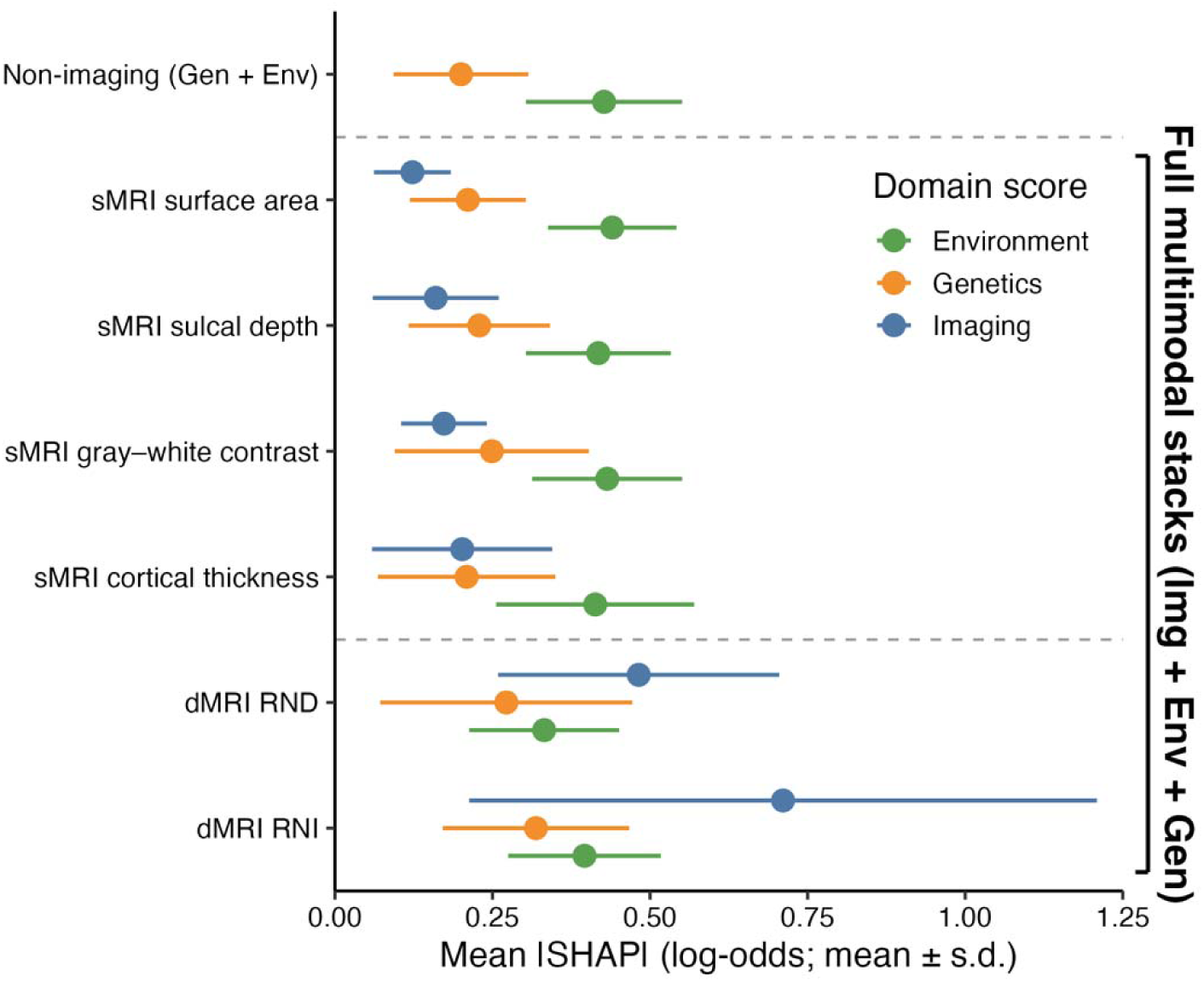
SHAP values illustrating domain contributions to multimodal stacks. Abbreviations: Env, Environmental domain; Gen, Genetic domain; Img, Imaging domain; sMRI, structural magnetic resonance imaging; dMRI, diffusion magnetic resonance imaging; PC, principal component; RND, restricted normalized directional diffusion; RNI, restricted normalized isotropic diffusion; SHAP, Shapley Additive exPlanations.

Stacked models built on the voxel-wise dMRI measures inverted this ordering. In full multimodal models, the imaging score was the largest contributor for both RND (0.482 ± 0.223 vs ENV = 0.332 ± 0.119; GEN = 0.272 ± 0.200) and RNI (0.711 ± 0.498 vs ENV = 0.396 ± 0.121; GEN = 0.319 ± 0.148). Importantly, this greater attribution did not translate into better discrimination: the two dMRI-based full multimodal stacks had the lowest ROC-AUCs of any full multimodal configuration (0.684 ± 0.027 and 0.683 ± 0.016), and their attributions carried the widest repeat-level SDs (up to ± 0.498). Attribution magnitude therefore reflects the influence of a domain score on the predicted log-odds rather than its accuracy.

Finally, imaging-only stacks produced near-zero attributions (0.002-0.063; Supplementary Table 9), which was lowest for RND (0.002 ± 0.008) and highest for cortical thickness (0.063 ± 0.098). SHAP values per domain for all combinations of multimodal stacks, including two-domain stacks, are included in Supplementary Figure 2.

## DISCUSSION

We applied a multimodal machine-learning framework to predict distressing and persistent psychotic-like experiences in a large, population-based cohort of youth drawn from the ABCD Study. Across all imaging modalities and stacked model configurations, findings converged upon neighborhood socioeconomic disadvantage and cumulative early adversity as the most important predictive features for dpPLEs, consistently outperforming neuroimaging features and polygenic/familial risk. The full multimodal ensemble achieved ROC-AUCs of up to 0.696 (sMRI) and 0.684 (dMRI), representing meaningful discrimination above chance. Critically, the environmental module alone approached the performance of the full multimodal model (ROC-AUC ≈ 0.687), whereas adding neuroimaging and genetics yielded only modest incremental changes (ΔAUC ≈ -0.004 to 0.009), underscoring that childhood socioenvironmental context carries disproportionate predictive weight for dpPLE risk. These findings carry implications for both the conceptualization of the risk architecture of childhood psychosis-spectrum phenomena and the design of scalable early identification tools.

Our findings that environmental features contributing most strongly to prediction of adolescent dpPLEs are aligned with a large body of literature, including recent emphasis in the ABCD Study, that socioeconomic context and exposome-related factors are critical for shaping youth mental health^34,35,47,62^. The three environmental features that consistently ranked highest across all SHAP analyses (area deprivation index, drug-related neighborhood crime, and cumulative adverse life events) align closely with established theoretical and empirical accounts of environmental risk for psychosis-spectrum experiences. Neighborhood socioeconomic deprivation has been implicated in psychosis risk through proposed mechanisms of social fragmentation, chronic stress activation, and reduced access to protective social resources^63–65^. The prominence of area deprivation index is consistent with emerging evidence that area-level structural disadvantage is a meaningful correlate of dpPLEs in the ABCD cohort^38,66,67^, and aligns with a recent meta-analysis demonstrating that neighborhood deprivation is associated with a 79% increase in rates of psychotic disorder incidence^64^.

The stable predictive contribution of cumulative adverse life events extends prior work demonstrating that childhood adversity is among the most potent environmental antecedents of psychotic-like experiences in population-based youth samples^38^. These findings also converge with a recent predictive analysis of general psychopathology risk in the ABCD Study, which reported that caregiver-reported psychosocial measures, including adverse childhood experiences and family mental health history, outperformed neuroimaging features^68^. Although that study targeted broad psychopathology risk rather than dpPLEs, the convergence of findings across independent analyses and distinct outcomes strengthens the conclusion that environmental and psychosocial measures carry disproportionate predictive weight in child and adolescent mental health prediction.

Within the genetic and family-history predictor domain, a family history of psychosis and/or depression was the strongest contributor, with the cross-disorder P-factor PGS ranking third. This ordering suggests a transdiagnostic genetic liability dimension, where dpPLEs may be capturing broader risk for psychopathology rather than risk that is more specific to psychosis-related outcomes. Although family history measures were grouped within the genetic module, these features also capture shared environmental exposures within the family unit, which may partly explain their greater predictive weight relative to PGS in this sample.

The modest contribution of disorder-specific PGS (schizophrenia, depression, bipolar disorder) is consistent with previous evidence of these scores showing limited and inconsistent associations with PLEs and related phenotypes in general population samples^69,70^. In another large youth cohort (the Philadelphia Neurodevelopmental Cohort; N = 7,225), multi-PGS models classified psychosis spectrum symptoms only marginally better than chance, with any predictive signal driven predominantly by participants of European ancestry^71^. The limited portability of PGS derived from predominantly European discovery GWAS to participants of non-European ancestry is a well-recognized limitation^30^. Ancestry-stratified sensitivity analyses confirmed that genetic predictive performance was driven by European-ancestry participants (Supplementary Tables 9 and 10). Although we prioritized retention of as many participants as possible to reflect the ABCD sample’s diversity, it is acknowledged that interpretation of PGS prediction in participants with non-European ancestry remains limited. Multi-ancestry PGS approaches thus represent a critical direction for future work.

The interplay between genetic liability and neighborhood environment should also be considered. A recent study found a significant negative interaction between schizophrenia PGS and neighborhood exposome scores on dpPLEs, such that the association between adverse neighborhood environments and dpPLEs was strongest among those with lower genetic risk^72^. This suggests that neighborhood exposures may exert a dominant influence on dpPLEs in the absence of a strong genetic predisposition, potentially attenuating the observable main effect of PGS^72^. This intersectional framing helps contextualize our findings of combined environmental and genetic features (ENV + GEN: ROC-AUC ≈ 0.697), which offered substantially greater discriminative value than genetic features alone.

Across all imaging features and stacked configurations, the incremental change in ROC-AUC from adding neuroimaging to the non-imaging ensemble was modest (ΔAUC ≈ -0.014 to - 0.001). This pattern replicates recent findings^68^ that similarly reported limited predictive performance of neuroimaging measures for general psychopathology risk in ABCD. However, the neuroimaging patterns contributing to dpPLE prediction hold important information about prospective neuroanatomical signatures of dpPLEs. For example, while global cortical surface area and RND across white matter tracts were found to contribute most in identifying youth with dpPLEs, other neuroimaging modalities showed more regionally specific patterns. RNI patterns, reflecting distribution diffusion among cell bodies, highlighted the basal ganglia and adjacent white matter as the most predictive features, while sensorimotor regions characterized by higher intracortical myelin drove prediction of GWC features. The latter findings are consistent with prior work noting potential alterations in peri-cortical myelin within sensory-motor pathways in patients at various clinical stages of psychotic disorders^26,73^. Broader patterns of cortical thickness and subcortical morphometry in youth with dpPLEs have also been found to be similar to adult neuropsychiatric illness signatures^74^. Our findings specifically suggest that such neuroanatomical changes, including measures of tissue microstructure and peri-cortical myelin, may already precede onset of observable PLE symptoms.

These models were developed in a population-based, non-help-seeking cohort and are best understood as a framework for characterizing premorbid risk signatures and informing future prediction efforts. Given that dpPLEs capture an emerging, non-diagnostic phenotype in the ABCD Study sample rather than an established clinical outcome, the predictive performance reported here (with peak performance of an AUC ≈ 0.7) should not be benchmarked against diagnostic models or expected to mirror clinical performance thresholds. Although we leveraged up to four waves of data from the ABCD Study dataset, predictors were limited to cross-sectional baseline measures only. Longitudinal trajectories, particularly for neuroimaging features, may elucidate useful temporal and developmentally grounded factors for dpPLE prediction in future work. We also acknowledge that some of the environmental risk measures, particularly adverse life events, relied on retrospective caregiver-report, which could introduce potential recall and appraisal bias; however, our environmental predictive features were strengthened by the use of geocoded measures (e.g., ADI, neighborhood crime).

## CONCLUSION

Using a large multimodal and prospective machine-learning framework, we demonstrate that neighborhood socioeconomic disadvantage and cumulative early adversity in childhood are the primary measurable risk substrates for persistent distressing psychotic-like experiences in adolescence, and are further informed by biological risk factors through family psychiatric history, polygenic influences, and broad patterns of morphometric and microstructural neuroimaging. These findings converge with evidence from broader psychiatric prediction analyses in ABCD that psychosocial and contextual features carry disproportionate predictive weight relative to neuroimaging in population-based youth cohorts, and they reinforce the importance of prevention strategies that address the structural social determinants of mental health risk early in development. Longitudinal extension and external validation represent critical next steps toward translating these findings into actionable clinical and public health frameworks.

## Data Availability

Data used in the preparation of this article were obtained from the Adolescent Brain Cognitive Development (ABCD) Study (https://abcdstudy.org/) version 6.0, held in the NIH Brain Development Cohorts Data Sharing Platform (https://www.nbdc-datahub.org/), as described above. Polygenic scores were derived from publicly available Psychiatric Genomics Consortium (https://pgc.unc.edu/for-researchers/download-results/) and iPSYCH (https://ipsych.dk/en/research/publications) genome-wide association summary statistics.

## Software and Data Availability

## Software

Analyses were implemented in Python 3.13.2 using scikit-learn 1.6.1, the SHAP package 0.48.0, and NumPy 2.2.4, pandas 2.2.3, SciPy 1.15.2, joblib 1.4.2, and h5py 3.12.1. Figures were produced with Matplotlib 3.10.1 and seaborn 0.13.2. Mixed-effects residualisation of the imaging data was performed with the Fast Efficient Mixed-effects Analysis (FEMA) package v.2.3.0, distributed as part of the cmig_tools package (https://github.com/cmig-research-group/cmig_tools). FEMA was called from Python through MATLAB R2023a Update 4. GWAS summary statistics were formatted using the *cleansumstats* function (https://github.com/BioPsyk/cleansumstats). Polygenic scores were computed with PRScs (https://github.com/getian107/PRScs).

## Data and code availability

Data used in the preparation of this article were obtained from the Adolescent Brain Cognitive Development™ (ABCD) Study (https://abcdstudy.org/) version 6.0, held in the NIH Brain Development Cohorts Data Sharing Platform (https://www.nbdc-datahub.org/). This is a multisite, longitudinal study designed to recruit more than 10,000 children aged 9-10 and follow them over 10 years into early adulthood. The ABCD Study® is supported by the National Institutes of Health and additional federal partners under award numbers: U01DA041048, U01DA050989, U01DA051016, U01DA041022, U01DA051018, U01DA051037, U01DA050987, U01DA041174, U01DA041106, U01DA041117, U01DA041028, U01DA041134, U01DA050988, U01DA051039, U01DA041156, U01DA041025, U01DA041120, U01DA051038, U01DA041148, U01DA041093, U01DA041089, U24DA041123, U24DA041147. A full list of supporters is available at https://abcdstudy.org/federal-partners.html. ABCD Consortium investigators designed and implemented the study and/or provided data but did not necessarily participate in the analysis or writing of this report. This manuscript reflects the views of the authors and may not reflect the opinions or views of the NIH or ABCD Consortium investigators.

Polygenic scores were derived from publicly available Psychiatric Genomics Consortium (https://pgc.unc.edu/for-researchers/download-results/) and iPSYCH (https://ipsych.dk/en/research/publications) genome-wide association summary statistics.

Analysis code will be made available upon publication at https://github.com/bridge-lab-research.

