## Supplementary Material for "Multimodal Prediction of Distressing and Persistent Psychotic-Like Experiences in Adolescence across Environment, Genetics, and Neuroimaging"

##### **Participants**

### Of the 11,878 baseline ABCD participants, 2,748 lacked sufficient data to define PLE outcomes and were excluded, leaving 9,130 with a classifiable PLE trajectory. Of these, 1,731 were further excluded for having transient or non-distressing PLEs, yielding a final sample of 7,399 (322 dpPLE cases, 7,077 low-PLE controls). The sMRI sample comprised 7,089 participants (3,152 female, 3,937 male; mean age 9.97 ± 0.63 years), of whom 308 (4.3%) met criteria for dpPLEs. The dMRI sample comprised 6,436 participants (2,909 female, 3,554 male; mean age 9.98 ± 0.63 years), of whom 279 (4.3%) met criteria for dpPLEs. The full multimodal stacked model, which required complete data across all modalities, was estimated in 7,399 participants.

##### **Genomic Sampling, Quality Control, and Analysis**

ABCD genetic information was retrieved from both saliva and blood and was assayed using the Smokescreen™ Genotyping array^1,2^. To improve SNP overlap with the external GWAS summary statistics, the markers from the Smokescreen array were imputed using the TOPMED imputation server^3^. These imputed variants were fractional dosages that were converted to an integer number of alleles using a best-guess threshold of 0.9. Genetic principal components (PCs) were generated from ABCD genomic data using PC-Air^4^ with default settings. PLINK 2.0^5^, was used to restrict our target data to only autosomal variants with a minor allele frequency of at least 1% (0.01). Genotype files were separated into three groups based on ancestry structure in the whole sample, a sample of European-like ancestry, and a sample of non-European-like ancestry (see *Genomic Ancestry Calculation* below for more details). We also removed SNPs missing in greater than 5% (0.05) of samples or with a Hardy-Weinberg equilibrium test ^6^ p-value lower than 0.001.

##### **Genomic Ancestry Estimation**

Genetic continental ancestry was estimated using SNPweights^7^ and external genomic reference panels for African, East Asian, European^8^, and Indigenous North and South American^9,10^ Ancestry Populations. ABCD subjects were separately categorized as having European-like ancestry (EUR) if they had an inferred genetic ancestry of at least 80% (0.8) consistent with the European ancestry reference panel, and were otherwise categorized as having non-European-like ancestry. This resulted in 4,745 participants (n=152 dpPLE group) with predominantly European ancestry and 2,654 participants (n=170 dpPLE group) with non-European-like ancestry.

#### **External GWAS Summary Statistics**

We used external and openly available summary statistics from Genome-wide association studies (GWAS) of depression^11^, schizophrenia^12^, bipolar disorder^13^, and cross-disorder^14^. External summary statistics were quality controlled and aligned to genome build GRCh38 using the *cleansumstats* function (https://github.com/BioPsyk/cleansumstats). See Supplementary Table 2 for details of GWAS samples.

#### **Polygenic Score Generation**

Best practices for imputing polygenic scores (PGS) in the ABCD Study sample are based on previous work showing that the Bayesian polygenic scoring method, PRScs^15^, maximized variance explained while maintaining a reasonable computational load compared to other Bayesian or pruning and thresholding methods^16^. Briefly, posterior effect sizes were generated using PRScs using 10,000 MCMC iterations and 5,000 burn-in iterations, which was found to balance reproducibility and computational intensity^17^. All other parameters were left at their default. LD reference panels required for this analysis were based on data from 1000 Genomes Phase 3 and matched the ancestry of the summary statistics. Posterior effect sizes were converted to polygenic scores using PLINK 2.0^18^.

**Prediction Framework**

***Nested Cross-Validation***

Predictive performance in classifying dpPLE versus lowPLE participants was estimated with repeated, family-based nested cross-validation. The outer loop consisted of 20 repeats of 5-fold cross-validation (100 outer splits), generated with scikit-learn StratifiedGroupKFold and incrementing the random seed each repeat. Family identifiers were used as the grouping variable so that siblings were never split between training and test sets. Splits in which the test fold contained only one outcome class were discarded and replaced until 100 valid splits were obtained. Within each outer training fold, hyperparameters were selected in a 5-fold inner loop, and stacking meta-features were generated in an additional 5-fold loop, both also using StratifiedGroupKFold grouped on family ID. ROC-AUC and PR-AUC were computed in each held-out test fold, averaged across the five folds of each repeat, and reported as mean ± SD across the 20 repeats.

***Non-Imaging Predictor Residualization***

Ordinal survey items were converted to numeric scores assuming equal spacing between adjacent response categories, and item scores were averaged to form each environmental feature. Similar to prior work^19^, each non-imaging predictor was residualized for nuisance covariates within the cross-validation framework described in the main text. Within each outer training fold, non-imaging predictors were residualized for nuisance covariates using ordinary least squares regression fitted on training data only. PGS and family history features were residualized for age at baseline, sex, study site, and the first ten genetic principal components; environmental features were residualized for age at baseline, sex, and study site. Each predictor was residualized in a separate univariate regression, with covariates entered as numeric terms, and residuals were computed as the observed minus model-predicted values. Coefficients estimated in the training fold were applied to the held-out fold to obtain test-set residuals.

#### ***MRI Preprocessing Parameters***

#### Imaging predictors were preprocessed within the cross-validation framework described in the main text, with all parameters estimated on the training fold only and applied unchanged to the held-out test fold. Within each outer training fold, vertex-wise sMRI and voxel-wise dMRI maps were residualized using the generalized least-squares (GLS) estimator in the Fast and Efficient Mixed Effects Algorithm (FEMA), with family as a random effect. Covariates were baseline age (z-scored within fold), sex, MR scanner (serial number combined with software version), and the first ten genetic ancestry principal components. Residualized maps were then mean-centered and standardized, and PCA (randomized SVD solver) was fit separately for each feature map (for example, cortical thickness and surface area for sMRI, and RNI and RND for dMRI), retaining 15 components per map. Component loadings were saved for visualization.

#### ***Classifier Hyperparameter Grid***

#### Separate base models were fit for each imaging feature map and for the genetic and environmental domains. All base models and the meta-learner used scikit-learn LogisticRegression ((SAGA solver, fixed random seed) tuned with GridSearchCV over three penalty types: an L1 penalty with C ∈ {0.0001, 0.001, 0.01, 0.1, 1.0, 10.0}, an L2 penalty with the same range of C, and an elastic-net penalty with C ∈ {0.0001, 0.0003, 0.001, 0.003, 0.01, 0.03, 0.1, 0.3, 1.0, 3.0, 10.0, 100.0, 1000.0} and L1 ratio ∈ {0.1, 0.2, …, 0.9}. Class weighting was tuned jointly with the penalty and C, with options of no weighting, scikit-learn "balanced" weights, or upweighting the dpPLE class by a factor of 2, 3, 5, or 10 relative to the lowPLE class. Imaging base models and the meta-learner were selected on PR-AUC, and non-imaging base models on ROC-AUC. The best configuration was refit on the full training fold. Maximum iterations were 5,000 (imaging), 2,000 (non-imaging), and 3,000 (meta-learner).

***Stacked Ensemble Construction***

Imaging, genetic, and environmental information was combined by stacked generalization. Each base model returned a predicted probability of dpPLE membership per participant, and these probabilities (meta-features) were used as predictors in a regularized logistic regression meta-learner. Stacking allowed each domain to be tuned to its own penalty type and strength, given that each domain differed substantially in dimensionality (15 imaging principal components versus 8 genetic and 8 environmental predictors). Meta-features for training participants were out-of-fold predictions from the additional family-grouped 5-fold loop described above. Each base model was then refit on the full outer training fold, with its own hyperparameter search, to generate meta-features for the test fold. Because the genetic and environmental models require complete data, participants missing any predictor in a domain were assigned the training-fold mean prediction for that domain. The meta-learner was fit directly to the resulting predicted probabilities (one to three columns) without rescaling. Five configurations were evaluated on identical participants and test folds: imaging only, imaging plus genetic, imaging plus environmental, genetic plus environmental, and all three domains, with each stack including a single imaging feature map. The imaging-only configuration has a single meta-feature and effectively captured a recalibration of the imaging base model.

***SHAP Estimation***

Feature contributions were quantified with SHAP, a unified, game-theoretic framework that attributes a model’s prediction to its input features^20,21^. SHAP values were computed within each outer split for held-out participants, using shap.LinearExplainer (v0.48.0, interventional feature perturbation) with a background sample of up to 200 participants from the training fold. For linear models under this setting, SHAP values have an exact closed form (the coefficient multiplied by the feature's deviation from its background mean) and are expressed as contributions to the predicted log-odds of dpPLE versus low PLE group status. For imaging base models, SHAP values indexed the 15 principal components; for stacked models, they indexed each base-model prediction. Importance was the mean absolute SHAP value per feature in each test fold, averaged across the five folds of each repeat and summarized as mean ± SD across the 20 repeats.

**SUPPLEMENTARY TABLES**

| **Domain** | **Measure** | **Description** |
| --- | --- | --- |
| Genetic | Polygenic scores (PGS) | PGS for Schizophrenia, Bipolar Disorder, Depression, Cross-disorder |
|  | Family History (FHx) | FHx of Psychosis, Mania, Depression from Family History Assessment Module Screener |
| Environment | Adverse childhood events | Parent-reported sum of distressing PhenX Life events occurring before 9 years of age |
|  | Area deprivation index | National percentile score based on baseline residential address |
|  | Drug abuse exposure | Obtained from the Uniform Crime Report from FBI, based on baseline residence |
|  | Estimated poverty | Proportion of individuals living in poverty (-125% poverty level) in neighborhood |
|  | Financial adversity | Sum of 7 parent-rated items from demographics |
|  | Lead exposure risk | Estimated from geocoded baseline residential address & data from https://www.vox.com/a/lead-exposure-risk-map; also described in ^22^ |
|  | Neighborhood safety | Sum of 3 parent-rated items of perception of neighborhood safety |
|  | Residential stability | Number of years at baseline residence |
| Imaging | Structural MRI (5124 vertices) | cortical thickness, surface area, sulcal depth, gray-white contrast |
|  | Diffusion MRI (155,179 voxels) | restricted normalized isotropic and restricted normalized directional |

**Supplementary Table 1**. Complete list of predictor variables across all domains (genetic, family history, environmental, and neuroimaging) used in the multimodal models, with corresponding measures and derivations.

| **GWAS Trait** | **Reference** | **NTotal** | **NCases** | **NControl** |
| --- | --- | --- | --- | --- |
| Depression | Als et al., 2023^11^ | 1,035,760 | 294,322 | 741,438 |
| Schizophrenia | Trubetskoy et al., 2022^12^ | 275,250 | 55,365 | 219,885 |
| Bipolar Disorder | Mullins et al., 2021^13^ | 413,466 | 41,917 | 371,549 |
| Cross-disorder (p-factor) | Grotzinger et al., 2026^14^ | 2,168,621 |  |  |

**Supplementary Table 2**. Details on GWAS summary statistics included for calculation of polygenic scores.

| **Model** | **ROC-AUC** | **PR-AUC** |
| --- | --- | --- |
| Environment only | 0.687 ± 0.003 | 0.091 ± 0.001 |
| Genetics only | 0.619 ± 0.020 | 0.080 ± 0.006 |
| Environment + Genetics | 0.697 ± 0.007 | 0.100 ± 0.006 |

**Supplementary Table 3**. Non-imaging domain prediction performance. ROC-AUC and PR-AUC are reported as mean ± SD across 20 repeat-level estimates, where each estimate is the average of 5 cross-validation folds within a single repeat for non-imaging predictors. Corresponds to Figure 2B in the main manuscript.

| **Model** | **European-like ancestry AUC**  (n_dpPLE_=152; n_lowPLE_=4593) | **Non-European-like ancestry AUC**  (n_dpPLE_=170; n_lowPLE_=2484) | **ΔAUC (95% CI)** |
| --- | --- | --- | --- |
| Environment | 0.712 ± 0.044 | 0.593 ± 0.046 | 0.119 (0.107–0.132) |
| Genetic | 0.679 ± 0.054 | 0.573 ± 0.050 | 0.106 (0.091–0.120) |

**Supplementary Table 4**. Ancestry-stratified sensitivity analysis: single-domain ROC-AUC for the environmental and genetic base models, estimated separately within participants of predominantly European-like ancestry compared non-European-like ancestry, with the between-ancestry difference in AUC (ΔAUC) and its 95% confidence interval.

| **Modality** | **ROC-AUC** | **PR-AUC** |
| --- | --- | --- |
| sMRI area | 0.619 ± 0.006 | 0.074 ± 0.002 |
| sMRI sulc | 0.599 ± 0.011 | 0.065 ± 0.003 |
| sMRI gwc | 0.595 ± 0.010 | 0.068 ± 0.002 |
| sMRI thk | 0.588 ± 0.008 | 0.072 ± 0.004 |
| dMRI rnd | 0.650 ± 0.005 | 0.087 ± 0.004 |
| dMRI rni | 0.611 ± 0.006 | 0.082 ± 0.003 |

**Supplementary Table 5**. Single-domain imaging prediction performance. ROC-AUC and PR-AUC are reported as mean ± SD across 20 repeat-level estimates, where each estimate is the average of 5 cross-validation folds within a single repeat for imaging predictors. Corresponds to Figure 2B in the main manuscript. *Abbreviations*. *sMRI, structural magnetic resonance imaging; dMRI, diffusion magnetic resonance imaging; sulc, sulcal depth; gwc, gray-white contrast; thk, thickness; rnd, restricted normalized directional diffusion; rni, restricted normalized isotropic diffusion.*

| **Modality** | **Model** | **ROC-AUC** | **PR-AUC** |
| --- | --- | --- | --- |
| sMRI area | Img + Env | 0.684 ± 0.021 | 0.092 ± 0.005 |
| sMRI area | Img + Gen | 0.586 ± 0.049 | 0.073 ± 0.012 |
| sMRI area | Img + Env + Gen | 0.695 ± 0.012 | 0.099 ± 0.007 |
| sMRI sulc | Img + Env | 0.678 ± 0.023 | 0.090 ± 0.005 |
| sMRI sulc | Img + Gen | 0.601 ± 0.042 | 0.074 ± 0.009 |
| sMRI sulc | Img + Env + Gen | 0.693 ± 0.016 | 0.096 ± 0.006 |
| sMRI t1_gwc | Img + Env | 0.688 ± 0.008 | 0.093 ± 0.005 |
| sMRI t1_gwc | Img + Gen | 0.617 ± 0.036 | 0.078 ± 0.008 |
| sMRI t1_gwc | Img + Env + Gen | 0.696 ± 0.010 | 0.099 ± 0.006 |
| sMRI thk | Img + Env | 0.681 ± 0.020 | 0.091 ± 0.006 |
| sMRI thk | Img + Gen | 0.611 ± 0.028 | 0.080 ± 0.007 |
| sMRI thk | Img + Env + Gen | 0.691 ± 0.019 | 0.097 ± 0.005 |
| dMRI rnd | Img + Env | 0.664 ± 0.035 | 0.091 ± 0.006 |
| dMRI rnd | Img + Gen | 0.603 ± 0.043 | 0.080 ± 0.011 |
| dMRI rnd | Img + Env + Gen | 0.684 ± 0.027 | 0.098 ± 0.007 |
| dMRI rni | Img + Env | 0.661 ± 0.030 | 0.094 ± 0.009 |
| dMRI rni | Img + Gen | 0.613 ± 0.027 | 0.084 ± 0.010 |
| dMRI rni | Img + Env + Gen | 0.683 ± 0.016 | 0.101 ± 0.007 |

**Supplementary Table 6**. Stacked ensemble prediction performance by imaging feature and stack configuration. ROC-AUC and PR-AUC are reported as mean ± SD across 20 repeat-level estimates, where each estimate is the average of 5 cross-validation folds within a single repeat. Corresponds to Figure 2B in the main manuscript. *Abbreviations*. *Env, environmental domain; Gen, genetic domain; Img, imaging domains; sMRI, structural magnetic resonance imaging; dMRI, diffusion magnetic resonance imaging; sulc, sulcal depth; gwc, gray-white contrast; thk, thickness; rnd, restricted normalized directional diffusion; rni, restricted normalized isotropic diffusion.*

| **Model** | **Top 3 features (mean SHAP ±SD)** |
| --- | --- |
| Env only | Area deprivation index percentile (0.193 ±0.013); Drug abuse violation total (0.161 ±0.013); Adverse Events (0.144 ±0.005) |
| Gen only | 1^o^ Family history psychiatric disorder (0.253 ±0.071); 1^o^ Family history depression (0.220 ±0.042); P-factor PGS (0.217 ±0.037) |
| Env + Gen | Env (0.427 ±0.124); Gen (0.200 ±0.107) |

**Supplementary Table 7**. Top-ranked features by SHAP importance for non-imaging models. Values are mean absolute SHAP, in log-odds units, reported as mean ± SD across 20 repeat-level estimates, where each estimate is the average of 5 cross-validation folds within a single repeat. Corresponds to Figure 3 in the main manuscript. See Supplementary Table 1 for full description of features. Abbreviations. *Env, environmental domain; Gen, genetic domain; SHAP, SHapley Additive exPlanations.*

| **Modality** | **Top 3 features (mean SHAP ±SD)** |
| --- | --- |
| sMRI area | PC1 (0.464 ±0.538); PC15 (0.076 ±0.050); PC10 (0.049 ±0.028) |
| sMRI sulc | PC1 (0.276 ±0.225); PC6 (0.163 ±0.180); PC11 (0.159 ±0.196) |
| sMRI gwc | PC2 (0.157 ±0.046); PC13 (0.147 ±0.038); PC5 (0.121 ±0.013) |
| sMRI thk | PC1 (0.223 ±0.316); PC14 (0.147 ±0.033); PC12 (0.103 ±0.031) |
| dMRI rnd | PC3 (3.858 ±0.434); PC11 (2.116 ±0.222); PC9 (2.054 ±0.127) |
| dMRI rni | PC9 (2.110 ±0.215); PC4 (1.667 ±0.167); PC7 (1.029 ±0.114) |

**Supplementary Table 8**. Top-ranked features by SHAP importance for imaging-only models. Values are mean absolute SHAP, in log-odds units, reported as mean ± SD across 20 repeat-level estimates, where each estimate is the average of 5 cross-validation folds within a single repeat. Corresponds to Figures 4 and 5 in the main manuscript, for voxelwise dMRI and vertexwise sMRI input features, respectively. Within each fold, SHAP values were computed for all held-out participants using an interventional linear explainer against a background sample of up to 200 training participants, then averaged in absolute value across participants. Absolute values index the magnitude of a feature's contribution to the predicted log-odds. *Abbreviations. SHAP, SHapley Additive exPlanations. sMRI, structural magnetic resonance imaging; dMRI, diffusion magnetic resonance imaging; sulc, sulcal depth; gwc, gray-white contrast; thk, thickness; rnd, restricted normalized directional diffusion; rni, restricted normalized isotropic diffusion.* *PC*. *principal component*.

| **Modality** | **Model** | **Domain contributions (mean \|SHAP\| ±SD)** |
| --- | --- | --- |
| sMRI area | Img only | sMRI area (Img) (0.010 ±0.019) |
| sMRI area | Img + Env | Env (0.470 ±0.114); sMRI area (Img) (0.143 ±0.071) |
| sMRI area | Img + Gen | Gen (0.283 ±0.138); sMRI area (Img) (0.167 ±0.052) |
| sMRI area | Img + Env + Gen | Env (0.440 ±0.102); Gen (0.211 ±0.092); sMRI area (Img) (0.123 ±0.061) |
| sMRI sulc | Img only | sMRI sulc (Img) (0.038 ±0.039) |
| sMRI sulc | Img + Env | Env (0.474 ±0.114); sMRI sulc (Img) (0.180 ±0.136) |
| sMRI sulc | Img + Gen | Gen (0.260 ±0.084); sMRI sulc (Img) (0.198 ±0.097) |
| sMRI sulc | Img + Env + Gen | Een (0.418 ±0.115); Gen (0.229 ±0.112); sMRI sulc (Img) (0.160 ±0.100) |
| sMRI gwc | Img only | sMRI gwc (Img) (0.027 ±0.031) |
| sMRI gwc | Img + Env | Env (0.400 ±0.124); sMRI gwc (Img) (0.153 ±0.081) |
| sMRI gwc | Img + Gen | Gen (0.266 ±0.085); sMRI gwc (Img) (0.216 ±0.076) |
| sMRI gwc | Img + Env + Gen | Env (0.432 ±0.119); Gen (0.249 ±0.154); sMRI gwc (Img) (0.173 ±0.068) |
| sMRI thk | Img only | sMRI thk (Img) (0.063 ±0.098) |
| sMRI thk | Img + Env | Env (0.414 ±0.120); sMRI thk (Img) (0.157 ±0.107) |
| sMRI thk | Img + Gen | sMRI thk (Img) (0.267 ±0.151); Gen (0.237 ±0.075) |
| sMRI thk | Img + Env + Gen | Env (0.413 ±0.157); Gen (0.209 ±0.141); sMRI thk (Img) (0.202 ±0.143) |
| dMRI rnd | Img only | dMRI rnd (Img) (0.002 ±0.008) |
| dMRI rnd | Img + Env | dMRI rnd (Img) (0.423 ±0.193); Env (0.284 ±0.129) |
| dMRI rnd | Img + Gen | dMRI rnd (Img) (0.560 ±0.240); Gen (0.269 ±0.247) |
| dMRI rnd | Img + Env + Gen | dMRI rnd (Img) (0.482 ±0.223); Env (0.332 ±0.119); Gen (0.272 ±0.200) |
| dMRI rni | Img only | dMRI rni (Img) (0.027 ±0.048) |
| dMRI rni | Img + Env | dMRI rni (Img) (0.883 ±0.798); Env (0.404 ±0.130) |
| dMRI rni | Img + Gen | dMRI rni (Img) (0.814 ±0.493); Gen (0.295 ±0.126) |
| dMRI rni | Img + Env + Gen | dMRI rni (Img) (0.711 ±0.498); Env (0.396 ±0.121); Gen (0.319 ±0.148) |

**Supplementary Table 9**. Top-ranked contributors by SHAP importance for stacked ensembles, by imaging feature and stack configuration. Values are mean absolute SHAP, in log-odds units, reported as mean ± SD across 20 repeat-level estimates, where each estimate is the average of 5 cross-validation folds within a single repeat. See main manuscript Figure 6. Features are the out-of-fold domain scores entering each stacker rather than raw measures; thus, values quantify each domain's contribution to the ensemble's predicted log-odds. *Abbreviations. SHAP, SHapley Additive exPlanations. Env, environmental domain; Gen, genetic domain; Img, imaging domain; sMRI, structural magnetic resonance imaging; dMRI, diffusion magnetic resonance imaging; sulc, sulcal depth; gwc, gray-white contrast; thk, thickness; rnd, restricted normalized directional diffusion; rni, restricted normalized isotropic diffusion.*

**SUPPLEMENTARY FIGURES**

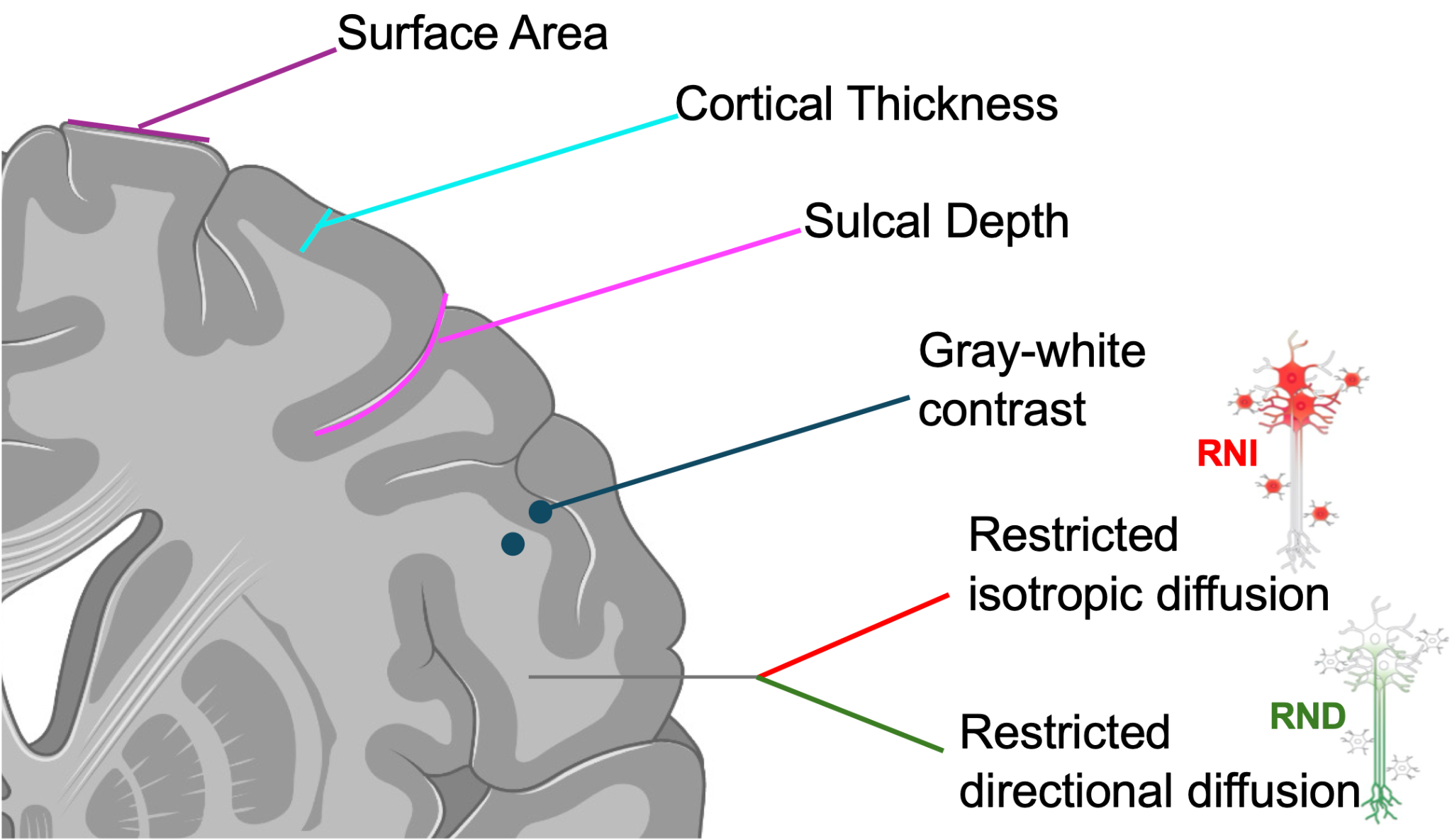

**Supplementary Figure 1**. Schematic of structural and diffusion MRI-derived neuroimaging features used as predictors, including cortical surface area, cortical thickness, sulcal depth, and gray-white contrast from structural MRI, and restricted normalized isotropic (RNI) and restricted normalized directional (RND) diffusion signal fractions from restriction spectrum imaging.

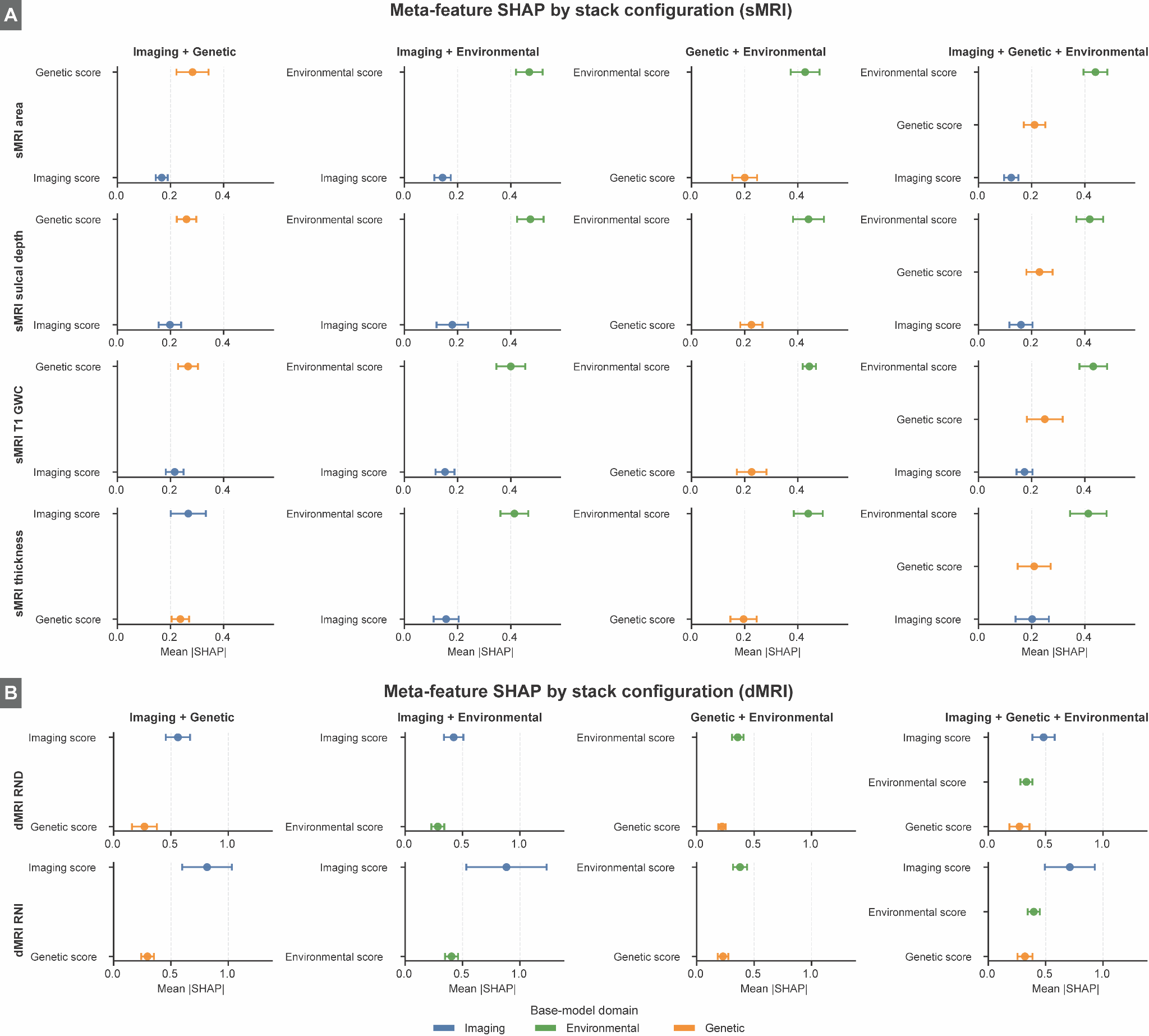

**Supplementary Figure 2**. SHAP values for all tested multimodal stack configurations and per domain contributions. *Abbreviations. SHAP, SHapley Additive exPlanations. sMRI, structural magnetic resonance imaging; dMRI, diffusion magnetic resonance imaging; GWC, gray-white contrast; rnd, restricted normalized directional diffusion; rni, restricted normalized isotropic diffusion.*
